# Novel application of capture-recapture methods to estimate the completeness of contact tracing during a large outbreak of Ebola Virus Disease, Democratic Republic of Congo, 2018-2020

**DOI:** 10.1101/2021.05.18.21257407

**Authors:** Jonathan A. Polonsky, Dankmar Böhning, Mory Keita, Steve Ahuka-Mundeke, Justus Nsio-Mbeta, Aaron Aruna Abedi, Mathias Mossoko, Janne Estill, Olivia Keiser, Laurent Kaiser, Zabulon Yoti, Patarawan Sangnawakij, Rattana Lerdsuwansri, Victor J. Del Rio Vilas

**Author notes:** Address for correspondence: Victor J. Del Rio Vilas, World Health Organization, South East Asia Regional Office, New Delhi, India.

## Abstract

Despite its critical role in containing outbreaks, the efficacy of contact tracing (CT), measured as the sensitivity of case detection, remains an elusive metric. We estimated the sensitivity of CT by applying unilist capture-recapture methods on data from the 2018-2020 outbreak of Ebola virus disease in the Democratic Republic of Congo. We applied different distributional assumptions to the zero-truncated count data to estimate the number of unobserved cases with a) any contacts and b) infected contacts, to compute CT sensitivity. Geometric distributions were the best fitting models. Our results indicate that CT efforts identified almost all (n=792, 99%) of the cases with any contacts, but only half (n=207, 48%) of the cases with infected contacts, suggesting that CT efforts performed well at identifying contacts during the listing stage, but performed poorly during the contact follow-up stage. We discuss extensions to our work and potential applications for the current COVID-19 pandemic.

## Introduction

Contact tracing (CT) is the process by which individuals who are believed to have come into contact with a confirmed case of an infectious disease during their infectious period are located and checked for the presence of the infection or disease. Under traditional approaches, CT involves three distinct steps; *contact identification*, in which potential contacts are identified through interview with the primary case; *contact listing*, in which those identified contacts are listed and communication established with them; and *contact follow-up*, in which those listed contacts are monitored for presence of infection or development of disease over a predefined period [1].

Due to its important role in case detection to monitor and curtail all chains of transmission, CT often forms part of the public health response to directly transmitted infectious diseases, both during the containment phase and as elimination of the disease from the population nears completion [2]. In recent years, CT has received widespread attention due to its critical role in the public health response to outbreaks of diphtheria [3], Ebola virus disease (EVD) [4–6], and the ongoing COVID-19 pandemic [7,8].

EVD is a disease caused by viruses of the genus Ebolavirus, family Filoviridae. Zoonotic spillover events from the animal reservoir have led to large, explosive outbreaks in West and Central Africa in recent years [9–12]. Owing to its high pathogenicity and virulence, an elimination control strategy is always adopted, aiming to ensure that all cases are promptly identified, isolated, and treated after disease onset (at which point cases become infectious), thereby limiting the opportunity for onward community spread. Although CT is a central pillar in the control strategy [13], there are currently no standard implementations of methods to assess a critical aspect of performance, its sensitivity, i.e. the ability to detect all the contacts and secondary infections resulting from detected cases.

One approach to quantifying this crucial metric is to employ capture-recapture (CRC) methods [14,15]. Broadly, this family of methodological approaches permits quantifying the number of individuals missing from lists, and subsequently estimate the sensitivity of the surveillance effort and the probability of case detection. While CRC has previously been used to estimate the number of unobserved cases of disease [16,17], such approaches typically rely on comparison of multiple lists, which are generally not available for lists of contacts. Therefore, we describe the application of a unilist capture-recapture approach [15] to quantifying the number of unobserved cases and contacts, and describe their likely sociodemographic profile, helping to identify risk factors that can be used to target limited resources at those unobserved cases most likely to generate onward transmission.

More precisely, we aim to address the following questions, from which we can derive CT sensitivity estimates:

1. How many cases with *any* contacts did CT miss?
2. How many cases with *infected* contacts did CT miss?

## Materials and methods

### Materials

During the course of the 2018-2020 EVD outbreak, a total of 3470 cases were detected across nine Health Zones (HZ) within three provinces. For our analyses, we included all cases and contacts identified by the Ebola Operational Centre (EOC) in Beni HZ, North Kivu Province, DRC between 31 July 2018 and 26 April 2020. Cases were confirmed according to World Health Organization (WHO)-recommended and national case definitions [18,19]. Cases were principally detected through three identification mechanisms: i) passive detection at healthcare facilities from clinically suspect individuals presenting symptoms consistent with EVD, ii) house-to-house active case finding by community health workers, and iii) tracing the contacts of probable and confirmed EVD cases. CT was coordinated by the Ministry of Public Health with support from WHO, and conducted by locally-recruited teams of contact tracers. Upon detection of a case, efforts to identify and list their contacts were undertaken.

For cases, our data contains information on basic sociodemographic characteristics (age, sex, Health Area of residence), and dates of disease onset and of isolation. For contacts, our data contains similar socio-demographic information, in addition to information on the daily follow up and final status of the contact (completed follow up, confirmed as EVD case, lost to follow up, never seen, and died during follow up).

### Methods

#### Exploratory data analysis

We presented the distribution of cases according to age, sex, and timing of disease onset. The distribution of the number of contacts per case were described between two distinct epidemic waves, with the Wilcoxon test used to explore differences in continuous variables and Chi-squared test used for categorical variables. “Superspreading”, or overdispersion in the offspring distribution of secondary cases arising from infectious individuals, may have profound impacts on control strategies in low-resource settings [20,21], and we describe the extent of this phenomenon in two ways: firstly by assessing the proportion of infectious individuals linked to 80% of onward transmission (an epidemiological extension of the Pareto principle which states that, for many outcomes, approximately 80% of the consequences arise from 20% of the causes [22]) using methods described by Endo *et al* [23]; and secondly by estimating the dispersion parameter (*k*) using methods described by Althaus [24].

A multivariable logistic regression model was used to explore risk factors associated with loss to follow-up, in which previously successfully traced contacts become untraceable at some point during the 21 days follow-up period. In such cases, contacts unable to be traced for three consecutive days are recorded as having been lost to follow-up, with no further attempts at tracing made.

#### Capture-recapture modelling

For each detected case, the CT process generates a list of individuals fitting the definition for a contact, some of whom may themselves have been infected and will eventually become secondary cases. From this, frequency distributions of cases with any contacts, and cases with contacts that resulted in secondary cases can be generated by first excluding (truncating) those cases with zero listed contacts. For example, the data can be binned into the number of cases with exactly one listed contact (*f*_1_), two listed contacts (*f*_2_), and so on to the number of cases with the maximum number of contacts (*f*_*m*_). Statistically, this leads to a zero-truncated observed count distribution of cases with at least one contact. By applying a unilist CRC approach designed to estimate unobserved population sizes using the distribution of count data within single lists [15], we can infer *f*_0_, the number of unobserved cases with at least one contact. In other words, we (1) assume that the number of *observed* contacts among cases who actually had contacts follows a parametric distribution, (2) find the best-fitting zero-truncated distribution based on the cases with at least one observed contact, and (3) use the expected number of cases with no observed contacts (calculated from the best-fitting distribution) as a proxy for the number of missed cases with contacts.

We take a generic zero-truncated variable *x* and consider the modelling of these zero-truncated count distributions. In particular, we are looking at the zero-truncated Poisson, Negative Binomial (NB) and geometric distributions (see Supplementary Methods).

If we distinguish between *n* observed and *f*0 unobserved cases, then an unbiased estimate of the unobserved number is given by the Horvitz-Thompson estimator, defined as

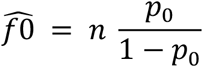

where *n* is the observed sample size of cases with at least one observed contact and *p*_0_ is the estimated probability of not observing a case with true contacts. We use maximum likelihood for model fitting, selecting the model with the smallest Akaike and Bayesian Information Criteria (AIC and BIC, respectively).

To estimate 95% confidence intervals (95%CIs), we use a parametric bootstrap, described as follows. Suppose that 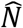 is the estimated size of the (observed and unobserved) population under a fitted model. Then, we generate *B* samples of size 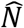 using the fitted model and its estimated parameter(s). For each sample, all zeros are truncated and the size estimate 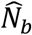 computed, for each of the samples *b=1*, …, *B*. We choose *B=10000* to minimise bootstrap simulation random error. The percentile method for constructing 95%CIs uses the 2.5^th^ percentile of the distribution of 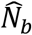 as the lower end and the 97.5^th^ percentile as the upper end.

## Results

### Exploratory data analysis

We identified 923 confirmed EVD cases in Beni HZ. A total of 80,556 contacts were listed through the CT process, of whom 6224 were duplicates, having been listed as the contact of more than one case, giving a total of 74,181 unique contacts to trace. The majority of cases for whom sex and age data were available were female (n = 515, 55.8%), while median (interquartile range, IQR) age was 25 (13-38) years. Among the total of 74,181 contacts listed through interviews with the case or their caregiver, 64,545 (87.0%) were successfully traced through contact tracing, leading to the detection of 396 secondary infections from the primary or index cases. Among those successfully traced, the median delay between last contact with the source case and first contact by the contact tracing teams was 4 days (interquartile range: 3-6 days).

The disease onset dates for the epidemic in Beni HZ spanned the period 31 July 2018 to 26 April 2020, and was bimodally distributed, with two waves peaking in October 2018 and again in June 2019 (Figure 1A). The second wave occurred during a period of insecurity in this active conflict zone that rendered response activities, including contact tracing, very difficult [25].

**Figure 1.**
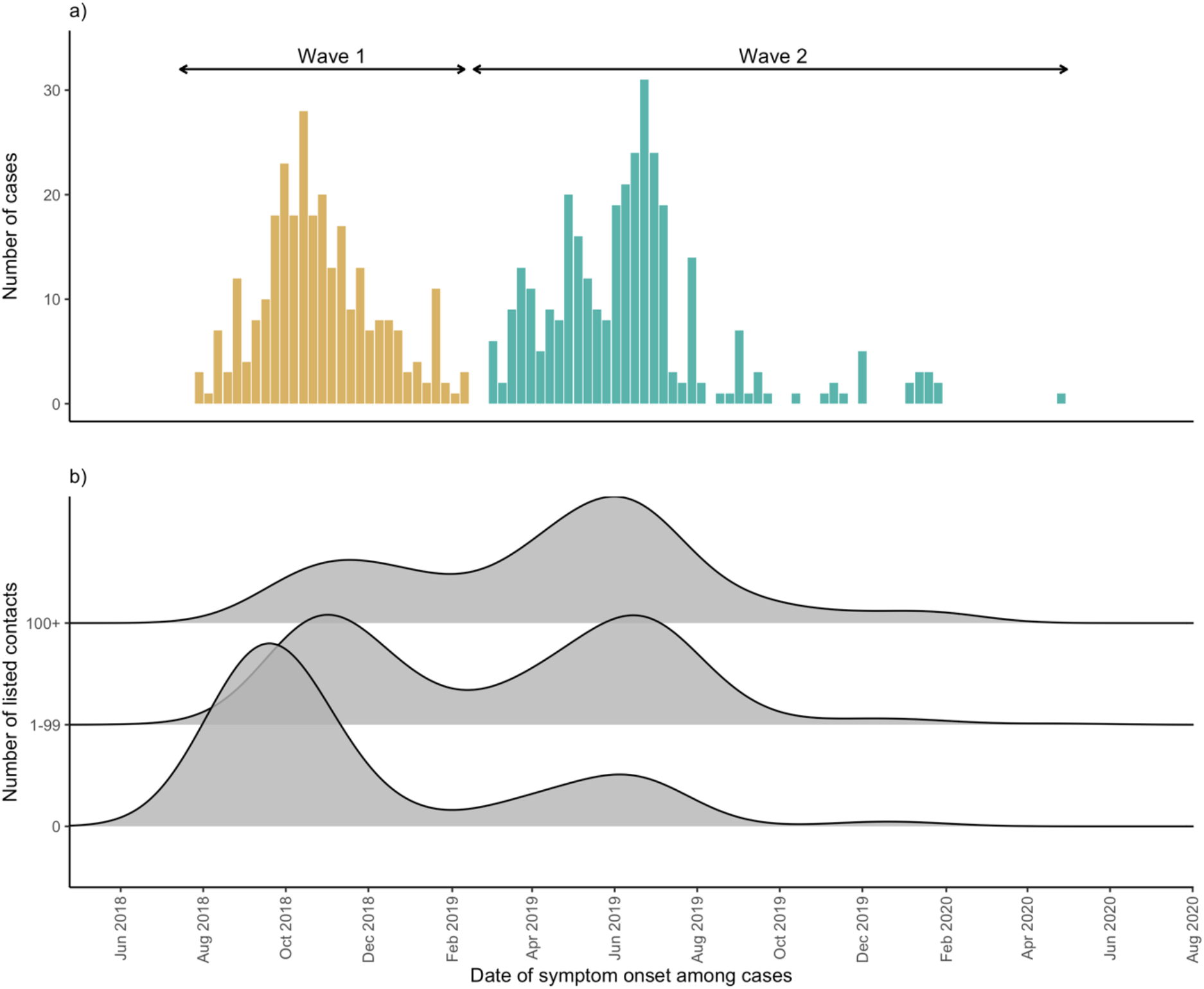
**a)** Epidemic curve by date of symptom onset among Ebola Virus Disease cases, Beni Health Zone, Democratic Republic of the Congo, 31 July 2018 - 26 April 2020. **b)** Distribution of dates of symptom onset among cases, by number of listed contacts. Data were smoothed using a non-parametric (Gaussian) kernel-based estimate, with automatic bandwidth selection (37.6 days).

The median (IQR) number of contacts per case was 61 (18 - 120) overall, but the Wilcoxon test showed that this was significantly lower during the first wave than the second (34 vs. 80, *p* < 0.001). Those cases with symptom onset during the first wave were more likely to have zero contacts listed than those with onset during the second wave (31.3% vs. 9.6%, *X*^2^ (1, *N* = 603) = 43.2, *p* < 0.001), while those with disease onset during the second wave were more likely to have a very large number (> 100) of listed contacts (Figure 1B).

Among the 923 cases recorded in Beni Health Zone, 131 (14.2%) cases had no listed contacts, while the remaining 792 (85.8%) reported at least one contact (Figures 2 and 3). Excluding those with no contacts identified, the median (IQR) and mean number of contacts was 74 (36 - 134) and 102, respectively, indicating a long-tailed distribution; indeed, the upper range was bound by a case with 823 contacts.

**Figure 2:**
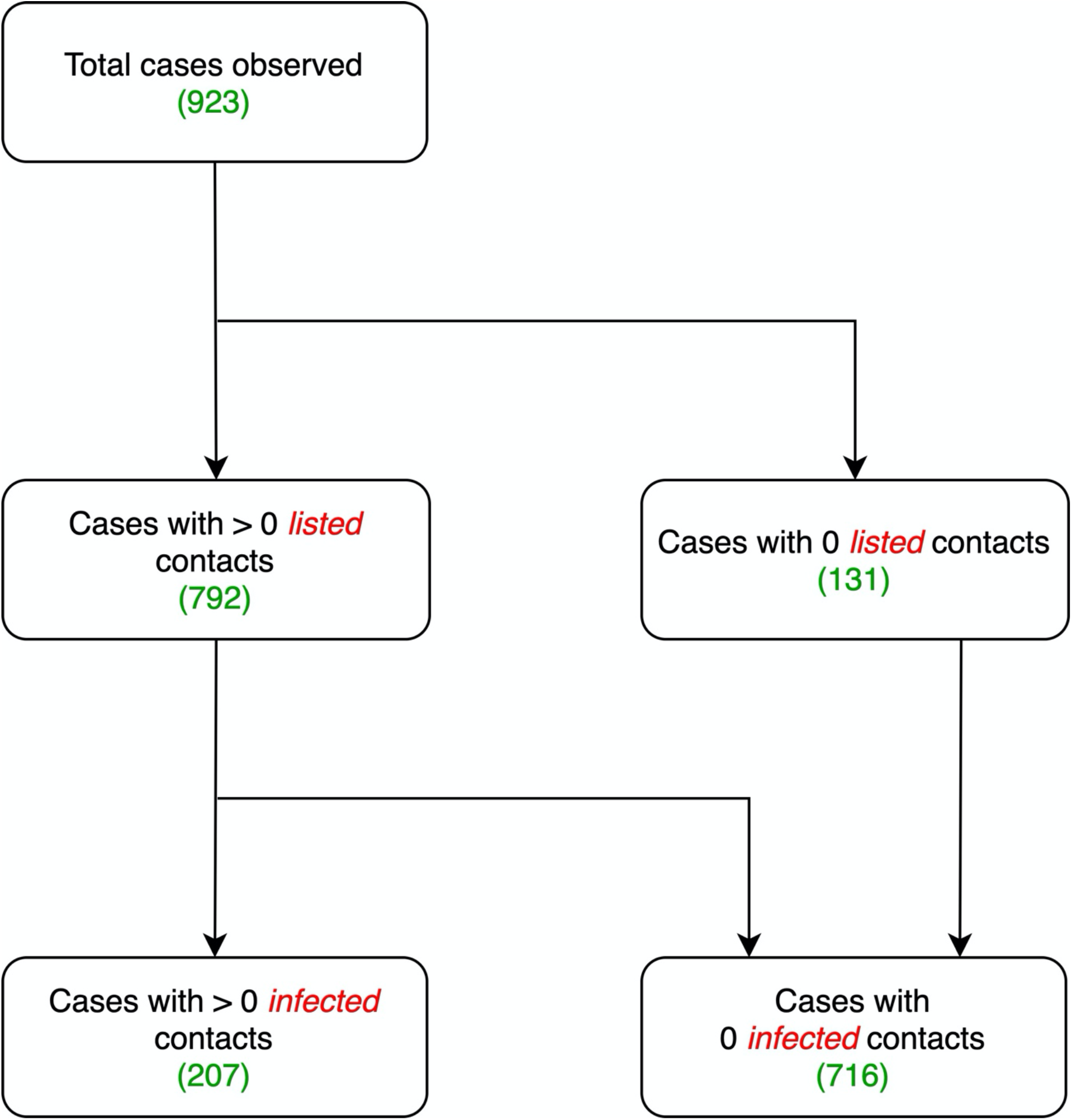
Flowchart showing breakdown of observed cases by number of listed and infected contacts among Ebola Virus Disease cases, Beni Health Zone, Democratic Republic of the Congo, 31 July 2018 - 26 April 2020.

**Figure 3:**
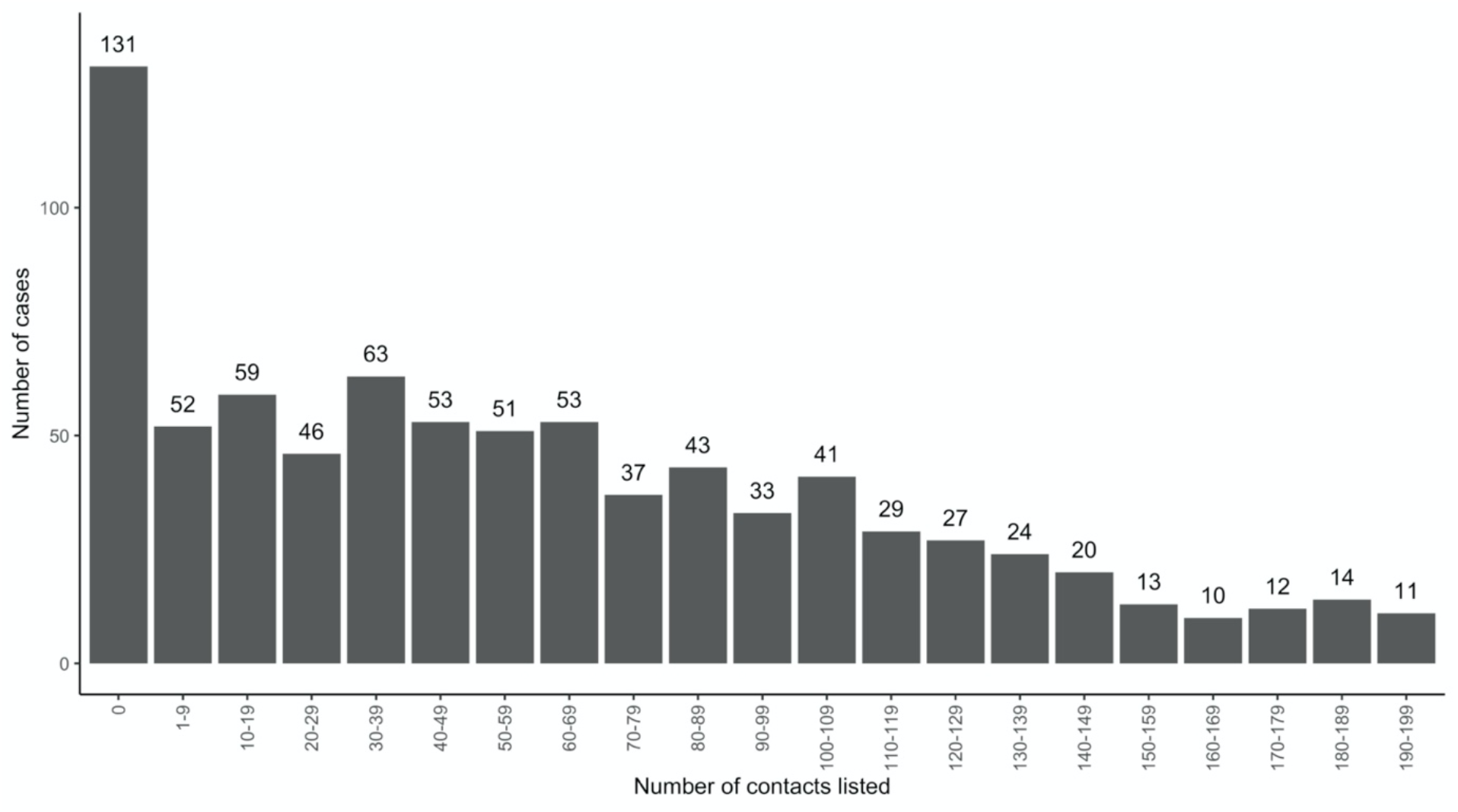
Frequency distribution of Ebola Virus Disease cases by number of listed contacts, Beni Health Zone, Democratic Republic of the Congo, 31 July 2018 - 26 April 2020. Note that the distribution is truncated to remove 101 cases with at least 200 contacts.

Among the 71,181 identified contacts, 64,545 (87.0%) were successfully traced, of whom 9222 (14.3%) were lost to follow up, 308 were recorded as testing positive for EVD, and 88 died, during the 21-day period of follow-up. For the purposes of our analyses, these 88 deceased contacts were assumed to have died of EVD due to the short time period between their contact with a confirmed EVD case and their death. Therefore, the total number of infected contacts was 396, or 0.6% of the total number of successfully traced contacts.

There was substantial overdispersion in the offspring distribution of secondary cases assumed to be caused by each infectious individual, with 80% of onward transmission linked to just 13.9% (95%CI 11.4 – 16.2) of primary cases, and *all* secondary infections concentrated among the contacts of approximately one quarter (207, 22.4%) of cases. Further, just 99 (10.7%) primary cases led to more than one secondary case (Figures 2 and 4). We estimated *k* as 0.27 (95%CI 0.20 – 0.33).

**Figure 4:**
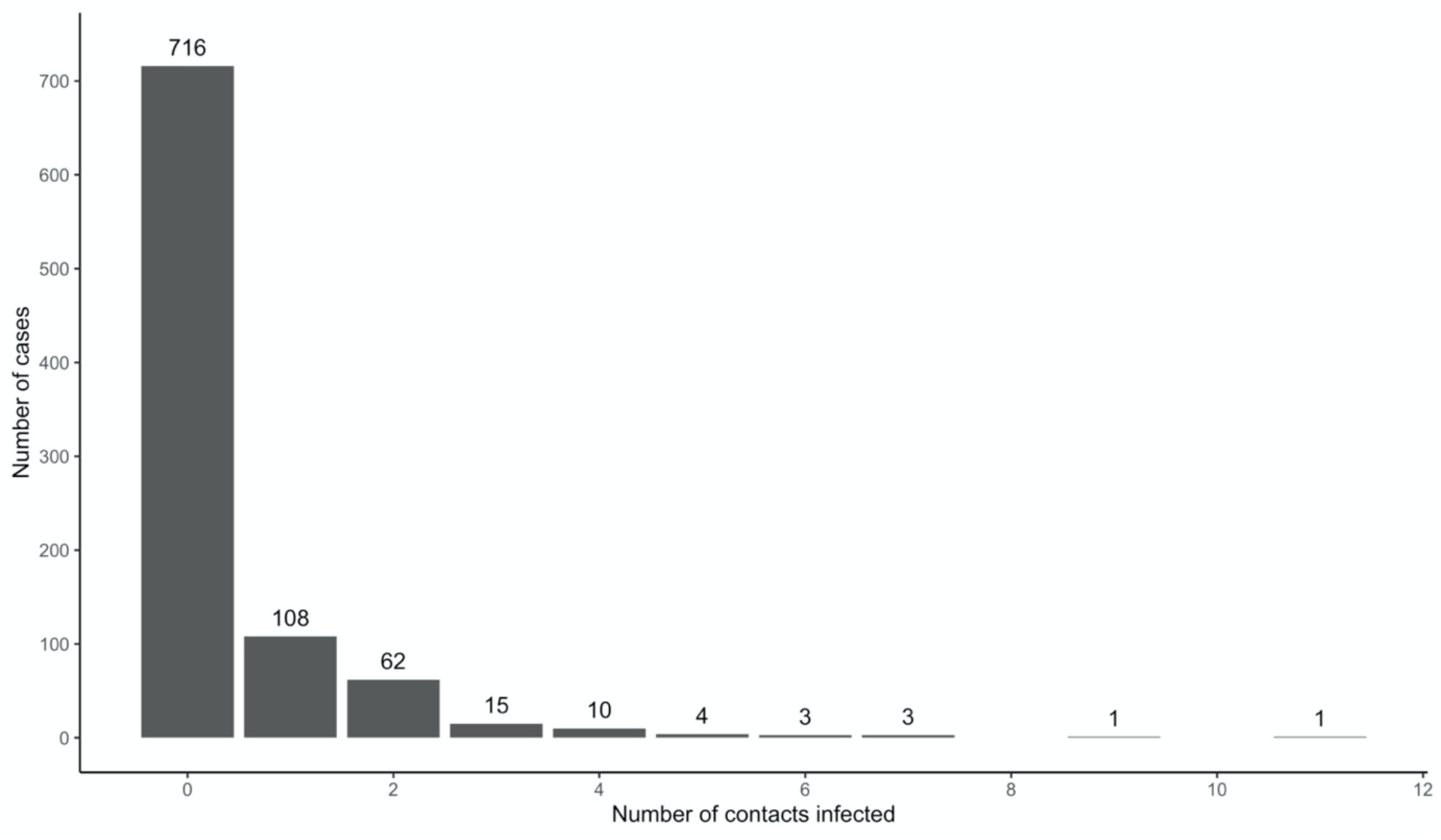
Frequency distribution of Ebola Virus Disease cases with infected contacts by number of infected contacts, Beni Health Zone, Democratic Republic of the Congo, 31 July 2018 - 26 April 2020.

In a multivariable logistic regression model, male contacts had slightly (but statistically significantly) greater odds of being lost to follow-up (odds ratio (OR) = 1.06, 95%CI 1.01 – 1.11, Table 1). Contacts in older age groups had significantly greater odds of being lost to follow up compared to contacts in the youngest age group (0-15 years), with the greatest effect being observed among those aged 60 years and older (OR = 1.65, 95%CI 1.47 – 1.86). Conversely, contacts traced during the second wave had lower odds of being lost to follow-up (OR = 0.83, 95%CI 0.79 – 0.88). Among cases with at least one infected contact, the median and mean number of infected contacts were 1 and 1.9, respectively.

**Table 1.** Multivariable logistic regression for predictors of loss to follow-up of contacts of Ebola Virus Disease cases, Beni Health Zone, Democratic Republic of the Congo, 31 July 2018 - 26 April 2020.

|  |  |  | Unadjusted |  | Adjusted |  |
| --- | --- | --- | --- | --- | --- | --- |
| Independent variable | Level | N | OR (95% CI) | p-value | OR (95% CI) | p-value |
| **Sex** | **Female** | 41349 | *Reference* | - | *Reference* | - |
|  | **Male** | 37296 | 1.07 (1.02 – 1.12) | **0.003** | 1.06 (1.01 – 1.11) | **0.013** |
| **Age group** | **0-14** | 20616 | *Reference* | - | *Reference* | - |
|  | **15-29** | 26142 | 1.18 (1.11 – 1.25) | **<0.001** | 1.19 (1.12 – 1.27) | **<0.001** |
|  | **30-44** | 17665 | 1.16 (1.09 – 1.24) | **<0.001** | 1.18 (1.10 – 1.26) | **<0.001** |
|  | **45-59** | 6157 | 1.56 (1.43 – 1.70) | **<0.001** | 1.55 (1.43 – 1.69) | **<0.001** |
|  | **60+** | 2599 | 1.64 (1.46 – 1.84) | **<0.001** | 1.65 (1.47 – 1.86) | **<0.001** |
| **Epidemic wave** | **First wave** | 14374 | *Reference* | - | *Reference* | - |
|  | **Second wave** | 66182 | 0.85 (0.81 – 0.90) | **<0.001** | 0.83 (0.79 – 0.88) | **<0.001** |

### Capture-recapture modelling

#### How complete was CT for all cases with at least one listed contact?

Among cases with at least one contact listed, the best fitting distribution of the count of cases with any contacts was given by the zero-truncated geometric model, which produced the lowest AIC and BIC (Table S1(a), supplementary materials). Note that this distribution was very long-tailed (Figure 5), indicating that the majority of cases with contacts were successfully detected, as with increasing mean of any count distribution, the probability for a zero count becomes small.

**Figure 5:**
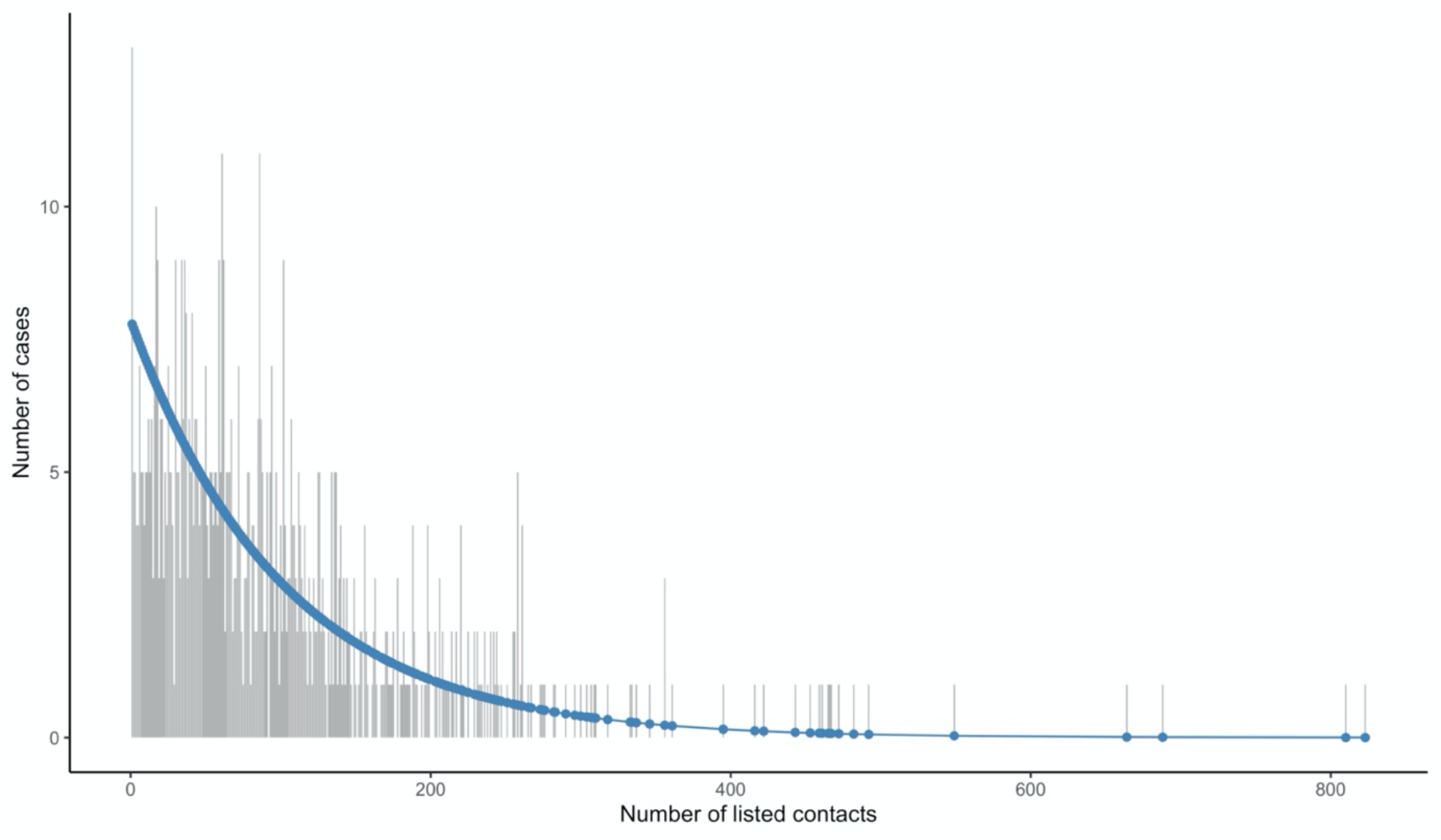
Observed and fitted (geometric) zero-truncated distribution of the total number of contacts for cases with *at least one contact listed*.

After fitting a zero-truncated geometric distribution, we used the Horvitz-Thompson estimator to estimate 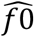 (the unobserved number of cases with any contacts) = 8 (95%CI = 8-10), where sample size (*n*) is 792 and *p*_0_(the estimated probability of not observing a case with at least one listed contact) 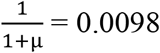 is with *µ* (the mean in geometric model) = 100.71). We used the parameterization 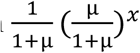 for the geometric, x=0,1,2,… Therefore, we estimated the sensitivity of contact tracing to detect cases with any contacts as 792/(792+8) = 0.99 (95%CI 0.98-0.99).

#### How complete was CT for cases with infected contacts?

Among cases with infected contacts, the best fitting distribution of count P was again given by the zero-truncated geometric model, which produced the lowest AIC and BIC (Table S1(b), supplementary materials). This distribution is concentrated on the lower counts from 1 to 4 (Figure 6), indicating that a substantial proportion of cases with infected contacts may not have been detected.

**Figure 6:**
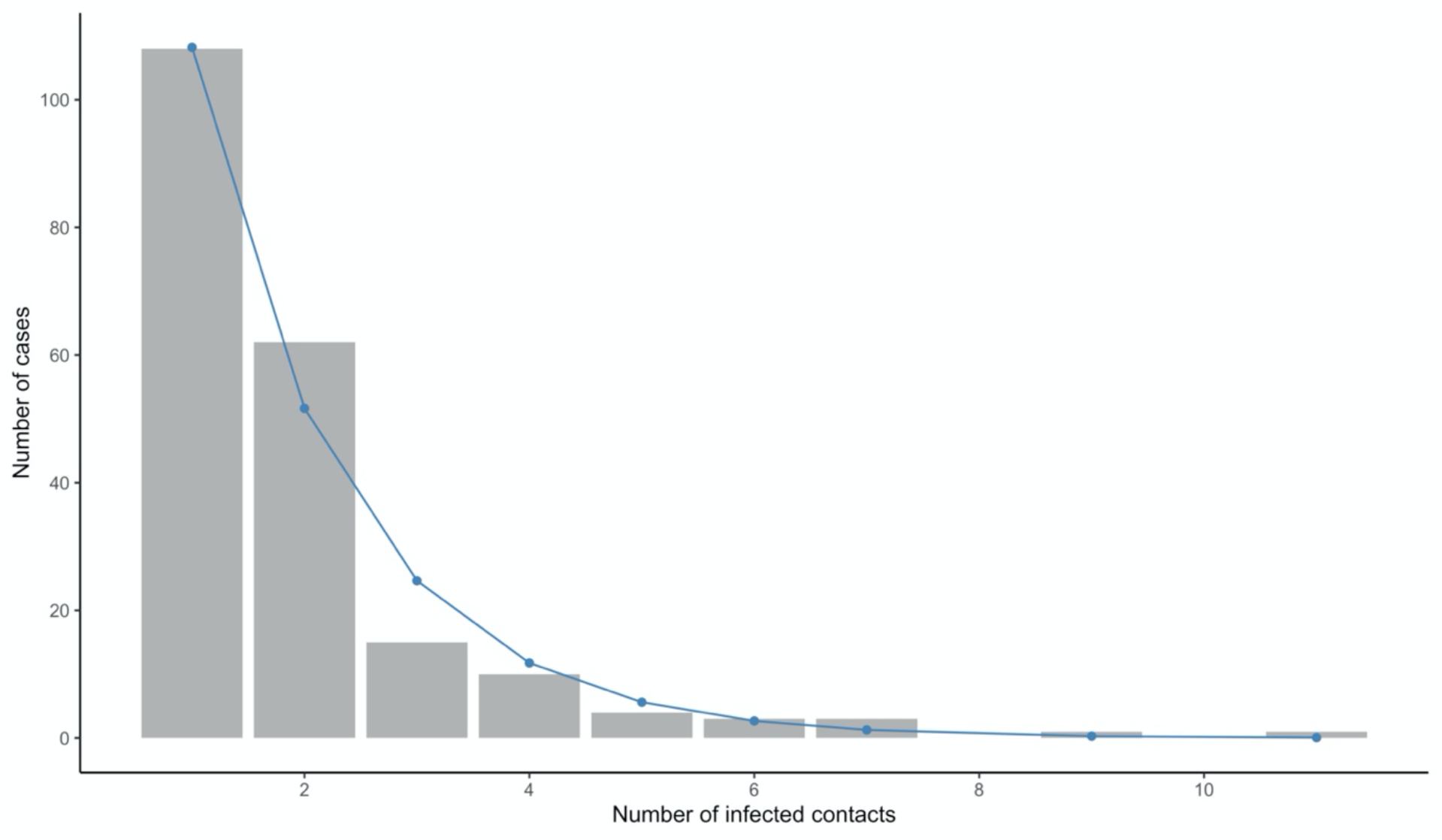
Observed and fitted (geometric) zero-truncated distribution of the total number of infected contacts for cases with *at least one infected contact listed*.

Using the Horvitz-Thompson estimator, we estimated 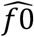 (the undetected number of cases with infected contacts) = 227 (95%CI = 171-241), where sample size (n) was 207 and *p*_0_ (the estimated probability of not observing a case with at least one *infected* contact) was 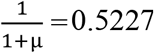, where μ = 0.9130. Therefore, we estimated the sensitivity of contact tracing to detect cases with infected contacts as 207/(207+227) = 0.48 (95%CI = 0.41-0.49).

The 792 cases with at least one listed contact (among whom we can be confident that at least a minimal investigation was conducted) can be grouped into three categories, according to the mean total number of contacts – those with no infected contacts identified, those with precisely one, and those with two or more (Table 2). Those cases with zero recorded infected contacts on average had fewer contacts overall, were slightly older, and slightly more likely to be female compared to the other groups.

**Table 2:** Distribution of cases with at least one listed contact according to number of infected contacts.

| # infected contacts | # cases | Mean (95%CI) # total contacts | Median age | % Female |
| --- | --- | --- | --- | --- |
| 0 | 585 | 85.7 (79.1 - 92.4) | 28.2 | 59.5 |
| 1 | 108 | 122 (102.0 - 141.0) | 23.6 | 56.7 |
| 2+ | 99 | 174 (144.0 - 204.0) | 25.6 | 54.6 |

## Discussion

Our findings suggest that CT efforts were broadly very successful at identifying cases with at least one contact, but much less successful at identifying cases with contacts who later develop symptoms. This is unsurprising, as the investigation component (typically by interview with cases under treatment and/or their caregivers) is much easier to conduct than the tracing component (typically requiring daily visits to a large number of difficult to locate and generally mobile individuals). This has important implications, as it is the contacts of these cases who have the potential to lead to ongoing chains of transmission when case investigation and contact tracing is inadequate, and in order to prioritise scarce resources, it is important to target those cases that lead to secondary infections among their contacts [20,21,26]. We report a high proportion of cases listing at least one contact (∼85%), compared to 27% and 44% during EVD outbreaks in Liberia [27] and Sierra Leone [26], suggesting that lessons about enhancing the quality of contact tracing from previous EVD outbreaks have been learned [4,5,26,27].

We found that those cases with infected contacts had a higher average number of overall contacts. This may be the result of three possible explanations. Firstly, individuals with more contacts are more likely to have at least one infected contact among these. Secondly, fewer overall listed contacts may be the result of poorly-conducted case investigations, which would be less likely to reveal the true extent of contacts made during the infectious period and therefore more likely to miss contacts, including those who had been infected. We found some evidence in support of this explanation, with the mean number of contacts increasing as the epidemic progressed, indicating a possible improvement in the quality of case investigation over time as staff became more accustomed to the procedure. Additionally, greater EVD awareness among the population may also have played an important role in better quality case investigations.

Thirdly, those cases with secondary infections among their contacts may qualitatively differ somehow from other cases. Cases with infected contacts were slightly younger and slightly more likely to be male, both of which are demographic factors that previously been shown to play an important role in the transmission patterns of EVD and other diseases. For example, prior evidence suggests that, in sub-Saharan Africa, younger individuals have more intense mixing patterns [28–30], while males are more likely to travel widely, which may lead to an increased likelihood of transmitting the disease to their contacts [30]. Indeed, a higher number of contacts overall suggests higher ‘node centrality’ within a given social network [31,32]. As those cases with more contacts and who are more active in social networks have been shown to play a greater role in disease transmission/more likely to be linked to confirmed cases among contacts, it is probable that, in this case, those cases with more contacts would be more likely to also have infected contacts within their social network. This is particularly true of diseases that demonstrate heterogeneous transmission, including EVD and COVID-19, and our results suggest a high degree of overdispersion and “superspreading”, in line with what has previously been reported during large EVD outbreaks [24]. Such overdispersion and clustering can lead to unexpected and explosive expansion, particularly among hidden chains of transmission, and one promising area of future research is to identify correlates of such overdispersion and use this information to target limited resources where they may have an outsized impact. Indeed, prior research suggests that, if highly infectious individuals can be predictively identified and targeted, the efficiency of control can be greatly enhanced, such that focussing half of all control effort on the most infectious 20% of cases can improve effectiveness up to threefold [20,21].

While it is possible to estimate the number of undetected cases with contacts, and those with infected contacts, it is not possible to identify whether these have been misclassified as having zero (infected) contacts or if they were undetected by the surveillance system in general. However, the greater probability of having zero contacts listed during the first epidemic wave suggests substantial misclassification in the period during which surveillance activities, including contact tracing, were being established and may have performed suboptimally, a finding reported for previous EVD outbreaks [4,26,27]. It is possible that some of the 131 cases listed as having no contacts were misclassified (Figure 2). Improved awareness of EVD symptoms and the potential for effective vaccination and treatment among the population may have played a role in improved ascertainment of contacts over time.

Loss to follow up of contacts is an important and well-documented concern during responses to EVD outbreaks, including during this outbreak with close to 15% of contacts not traced for the duration of the follow-up period. This may in part explain the misclassification of cases as having zero infected contacts if some of their contacts developed symptoms during the period in which they were not able to be directly observed.

We have described a novel application of CRC models to address an important question that has wide public health significance for the control of directly-transmissible diseases. The approach described is disease-agnostic, and is in principle applicable to other CT mechanisms, such as those for COVID-19, where the true number of COVID-19 infections has been estimated to be three to eight times larger than those reported [33]. Specifically for COVID-19, our approach provides a statistical framework to assess the significance of the increasing list of indicators to track CT performance. With data on resources invested in CT deployment (e.g. size of CT teams, time to successful tracking) our models can estimate efficiency, as the ratio of resources over efficacy as informed by CT sensitivity.

### Limitations

While the method described herein proposes a robust framework to assess the sensitivity of contact tracing, there are important limitations. There is no gold standard list of contacts against which to validate this method, but the method itself has been validated to estimate true population size in a variety of other settings [34]. The dataset does not permit the distinction between cases who were confirmed to have no contacts after a thorough case investigation and cases having no listed contacts due to no (or inadequate) case investigation. However, our method may in fact help to identify the magnitude of the misclassification arising from this. The inferences made are exclusively informed by the definition of cases as defined by CT protocols; for example, our results would not inform the sensitivity of CT as applied to asymptomatic EVD cases if these individuals are not part of the testing strategy. Finally, we have not adjusted for observed or unobserved heterogeneity, such as age, sex, profession, geographic location of the cases and delays within the contact tracing process, and further work is planned to incorporate such considerations.

## Conclusions

In conclusion, CT is crucial to the containment of certain disease outbreaks. However, as with any surveillance activity, CT suffers from underreporting and typically poor sensitivity. The consequences of poor ascertainment and misclassification can be disastrous in the containment stages of an outbreak, potentially creating explosive expansion among hidden chains of transmission, particularly during containment and de-escalation phases.

We have applied a novel method to estimating a crucial yet elusive key performance indicator of a crucial component of the public health response to epidemics, namely the sensitivity and efficacy of contact tracing, as applied to a recent outbreak of EVD. The method demonstrated that the majority of cases with any contacts were observed, suggesting that the case investigation component of CT performed well, while less than half of cases with infected contacts were observed, suggesting that the contact follow-up component of CT performed poorly in this setting.

This method can be extended to assess the efficacy of CT for any disease, including COVID-19, a disease for which CT has been identified as a crucial component of the response activities, yet for which key performance indicators and measures to assess these are yet to be established.

## Data Availability

The datasets analysed during the current study are not publicly available due to patient confidentiality concerns, but are available from the corresponding author on reasonable request.

## Acknowledgments

We thank the population for their participation conducting this study. We acknowledge the enormous dedication of the various organisations and individuals that responded and provided healthcare to this population and supported the public health response during this outbreak. The authors received no specific funding for this work. The authors alone are responsible for the views expressed in this article and they do not necessarily represent the views, decisions or policies of the institutions with which they are affiliated.

## Supplementary materials

### 1. Details of models used for modeling zero-truncated distributions

The probability mass function (pmf) of a *Poisson distribution* is given by:

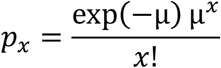

for *x* = 0,1,2,…, where μ is the parameter (mean) of the of the Poisson distribution.

The pmf of a *negative binomial distribution* is given by:

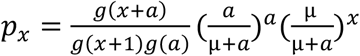

for *x* = 0,1,2,…, where *g*(.) is the Gamma function, and μ (mean) and *a* are the parameters of the negative binomial distribution.

The pmf of a *geometric distribution* is given by:

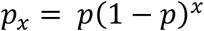

for *x* = 0,1,2,…, where *p* is the event parameter (between 0 and 1) in the geometric distribution. Note that

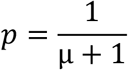

where μ is the mean of the geometric distribution.

The pmf’s of the associated zero-truncated distributions for all of the three distributions above are

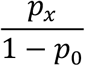

for *x* = 1,2,…

Note that the geometric arises as a special case of the negative binomial, where *a*=1 and

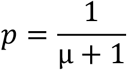

which also corresponds to *p*_0_.

**Table S1:** Model performance for a) count of cases with any contacts and b) count of cases with *infected* contacts

|  | a) Cases with any contacts |  | b) Cases with infected contacts |  |
| --- | --- | --- | --- | --- |
| Model | AIC | BIC | AIC | BIC |
| Poisson | 66635 | 66640 | 590 | 593 |
| Negative binomial | 8895 | 8905 | 551 | 558 |
| **Geometric** | **8900** | **8904** | **550** | **553** |

### 2. Details of methods used to estimate the unobserved frequency of counts

We have also considered other estimation methods such as the nonparametric estimator of Chao and the Turing estimator to estimate the unobserved frequency when count of observations is zero (*f*_0_):

MLE

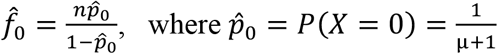

and μ = mean of geometric distribution.

Chao estimator

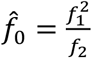

Turing estimator 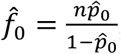, where 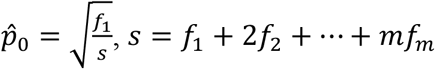, and *m* is the largest observed count.

The population size *N* is estimated by 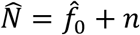, where *n* is the number of observed individuals. Note that we have based the analysis in our paper so far on the MLE. Table S2 presents the results for the indicator of interest for both the total population size as well as the number of unobserved individuals (cases). Table S3 provides uncertainty estimates for the values given in Table S3.

**Table S2:** Estimated population size and frequency of unobserved individuals using real datasets.

| | $\hat{N}_{mle}$ | $\hat{N}_{chao}$ | $\hat{N}_{turing}$ | $\hat{f}_{0mle}$ | $\hat{f}_{0chao}$ | $\hat{f}_{0turing}$ |
| --- | --- | --- | --- | --- | --- | --- |
| Contacts listed | 800 | 826 | 802 | 8 | 34 | 10 |
| Infected contacts<br>and contacts died | 434 | 395 | 433 | 227 | 188 | 226 |

**Table S3:** Estimated population size, unobserved frequency, and the 95% confidence interval using parametric bootstrap (pb) and bootstrap percentile (bp) methods

| | Bootstrap method for $\hat{N}$ | | | | | |
| --- | --- | --- | --- | --- | --- | --- |
| | $\hat{N}_{mle}$ | $\hat{N}_{chao}$ | $\hat{N}_{turing}$ | pb.CI.mle | pb.CI.chao | pb.CI.turing |
|  |  |  |  | bp.CI.mle | bp.CI.chao | bp.CI.turing |
| Contacts listed | 799.87 | 799.36 | 799.73 | 799.49-800.25<br>779.34-800.45 | 791.93-806.80<br>793-828 | 797.90-801.55<br>769.80-802.53 |
| Infected contacts<br>and contacts died | 434.59 | 435.48 | 434.55 | 400.47-468.72<br>390.81-491.46 | 365.29-505.68<br>350.46-568.78 | 397.99-471.10<br>385.97-495.90 |
| | Bootstrap method for $\hat{f}_0$ | | | | | |
| | $\hat{f}_{0mle}$ | $\hat{f}_{0chao}$ | $\hat{f}_{0turing}$ | pb.CI.mle | pb.CI.chao | pb.CI.turing |
|  |  |  |  | bp.CI.mle | bp.CI.chao | bp.CI.turing |
| Contacts listed | 7.87 | 7.36 | 7.73 | 7.49-8.25<br>7.34-8.45 | 0-14.80<br>1-36 | 5.90-9.55<br>4.80-10.53 |
| Infected contacts<br>and contacts died | 227.59 | 228.48 | 227.55 | 193.47-261.72<br>183.81-284.46 | 158.29-298.68<br>143.46-361.78 | 190.99-264.10<br>178.97-288.90 |

## Notes

### Competing Interest Statement

The authors have declared no competing interest.

### Author Declarations

Comité d'éthique de l'Ecole de Santé Publique de l'Université of Kinshasa (DRC) reference: ESP/CE/03/2021

